# Health-Weighted Delimitation: A Public Health-Informed Framework for Equitable Electoral Representation in India

**DOI:** 10.1101/2025.08.22.25334059

**Authors:** Karthik Balajee Laksham

## Abstract

**Background:** India’s upcoming parliamentary delimitation, based solely on population size, risks penalising states with successful public health and birth control policies amid concerns about reduced representation in the South. No previous study has integrated health metrics into electoral representation. The study developed and tested a health-weighted delimitation model that includes public health metrics to ensure equitable electoral representation while preserving the current allocation of seats.

**Methods:** Data for all 28 states and 8 UTs were compiled from the National Family Health Survey-5, Sample Registration System, and Census 2011. Five indicators, like projected population (2021), infant mortality rate (IMR), maternal mortality ratio (MMR), total fertility rate (TFR), and female literacy, were normalised. TFR, IMR, and MMR were reverse-scored so that higher values reflected better health performance. Principal component analysis (PCA), conducted using R software, generated a composite Health-Weighted Performance Score (HWPS), with the first principal component explaining 70.3% of the total variance. Seats were allocated in proportion to HWPS under three scenarios: the current allocation (543 seats), a 10% expansion (597 seats), and a 15% expansion (625 seats). The algorithm guaranteed that no region received fewer seats than in 2019 and capped increases at 10% or 15%. Robustness was assessed using 1,000 Monte Carlo simulations (+/-10% Gaussian noise) and +/-20% sensitivity analyses.

**Results:** The HWPS ranged from 1 (Bihar) to 100 (Lakshadweep), with higher scores in states with strong health and literacy outcomes. Under a 10% expansion, 26 of 36 regions gained seats, with none losing representation. Health-advanced Kerala increased from 20 (in 2019) to 22 seats (in a 597-seat scenario). Populous states like Bihar and Uttar Pradesh received modest increases. The Seat allocations remained consistent across simulations and sensitivity analyses (+/-1 seat in 95% simulations).

**Conclusions:** The health-weighted parliamentary seat allocation model is a novel and transparent framework that integrates public health into political representation. This model promotes electoral equity by maintaining current representation and rewarding health progress.

## Introduction

India’s population policy has evolved significantly. The forced sterilisation programmes, especially during the 1975-77 Emergency, caused public distrust and led to a shift toward voluntary methods [1]. The Reproductive and Child Health strategy (1997) encouraged decentralised population control. [1] The National Population Policy (2000) focused on maternal and child health to support stable population growth, shaping current demographic trends. However, regional differences persist. The birth rates are higher in northern states like Bihar and Uttar Pradesh (20.8-26.2 per 1,000 people) than in southern states like Tamil Nadu and Kerala (11.1-15.3 per 1,000) [1]. By 2036, India’s population is projected to reach 1.52 billion. Almost half of this growth will come from northern states, while southern states will account for just 9% [2]. These gaps require customised public health plans.

The Lok Sabha, the lower house of the Indian parliament, has 543 seats, fixed since 1976 based on the 1971 Census [3]. The 84th Amendment delayed seat reallocation until after 2026, using the first census post-2026, likely 2031 [3]. The upcoming delimitation will redraw electoral boundaries based on new census data, favouring populous states like Uttar Pradesh while potentially reducing representation for states like Kerala and Tamil Nadu, which controlled population growth [3]. On March 22, 2025, southern state leaders, including Tamil Nadu’s Chief Minister, called it a “sword of Damocles” and a “political attack” on states with strong health systems [4-7]. The Indian National Congress, the main opposition party in India, has warned that population-based delimitation could penalise states like Tamil Nadu and Kerala for successful population control, and possibly reduce their parliamentary seats [6].

Political representation shapes public health through its influence on resource allocation and political priorities. States with more seats in parliament receive more funding and support, while smaller states risk losing influence, which impedes health progress. The population-based approach could reduce the impact of healthy states on the federal balance and discourage investment in health. Linking health outcomes to seat allocation can reduce the equity disadvantage based on population alone. However, no previous research has explicitly linked health metrics to electoral seat allocation. This study proposes a health-weighted delimitation model to align electoral representation with public health achievements fairly.

### Health-Weighted Delimitation

This model uses health indicators like Total Fertility Rate (TFR), Infant Mortality Rate (IMR), Maternal Mortality Ratio (MMR), and female literacy. It protects states with weaker systems and rewards states for improving health, to make electoral representation more equitable. This idea is inspired by global examples, such as the EU’s degressive proportionality, which gives smaller states a balanced representation [18-20]. Amartya Sen’s Capabilities Approach views development as expanding freedoms to live healthy, meaningful lives [8]. Public Choice Theory suggests leaders prioritise self-interest, like votes or power [9]. These theories support a system in which countries gain political influence by improving health and creating a just and incentive-based framework. The model encourages countries to reduce TFR, IMR, and MMR and increase health investment. For example, the states that improve maternal and child health could gain more prominence or influence.

No country currently ties health metrics to electoral representation, but global models offer ideas. Bhutan’s Gross National Happiness index focuses on well-being [17], and the EU’s system supports smaller states [18-20]. The UN’s Human Development Index uses health and education to measure progress[24]. The World Health Organisation’s Equity Framework promotes equitable access to health services and is in line with the objectives of the model [25]. The United States bases its seats on population and adjusts for demographic changes. [21], while Australia and Canada take socioeconomic factors into account [22,23]. India has experience using data to improve health and education in underserved areas, such as through the NITI Aayog’s Aspirational Districts Programme. [16] With strong national data systems like NFHS and SRS, India is well-placed to lead the innovative approach to link political representation with health outcomes.

### Methodology

Data for all 28 Indian states and 8 Union Territories were collected from the latest publicly available sources, such as the National Family Health Survey-5 (NFHS-5), the Sample Registration System (SRS), and Census 2011. Five key indicators were taken for each region: the infant mortality rate (IMR per 1,000 live births), maternal mortality ratio (MMR per 100,000 live births), total fertility rate (TFR), female literacy rate (percentage) and the projected population for 2021. IMR, MMR, and TFR reflect maternal and child health, while female literacy influences health behaviours and outcomes. These indicators were chosen because they have a direct relationship with health equity. Table 1 summarises the five health indicators and their relevance to equitable representation. In all simulated scenarios, every state and Union Territory retained the number of Lok Sabha seats allocated in 2019 as a minimum guarantee (“no-loss” safeguard).

**Table 1:**
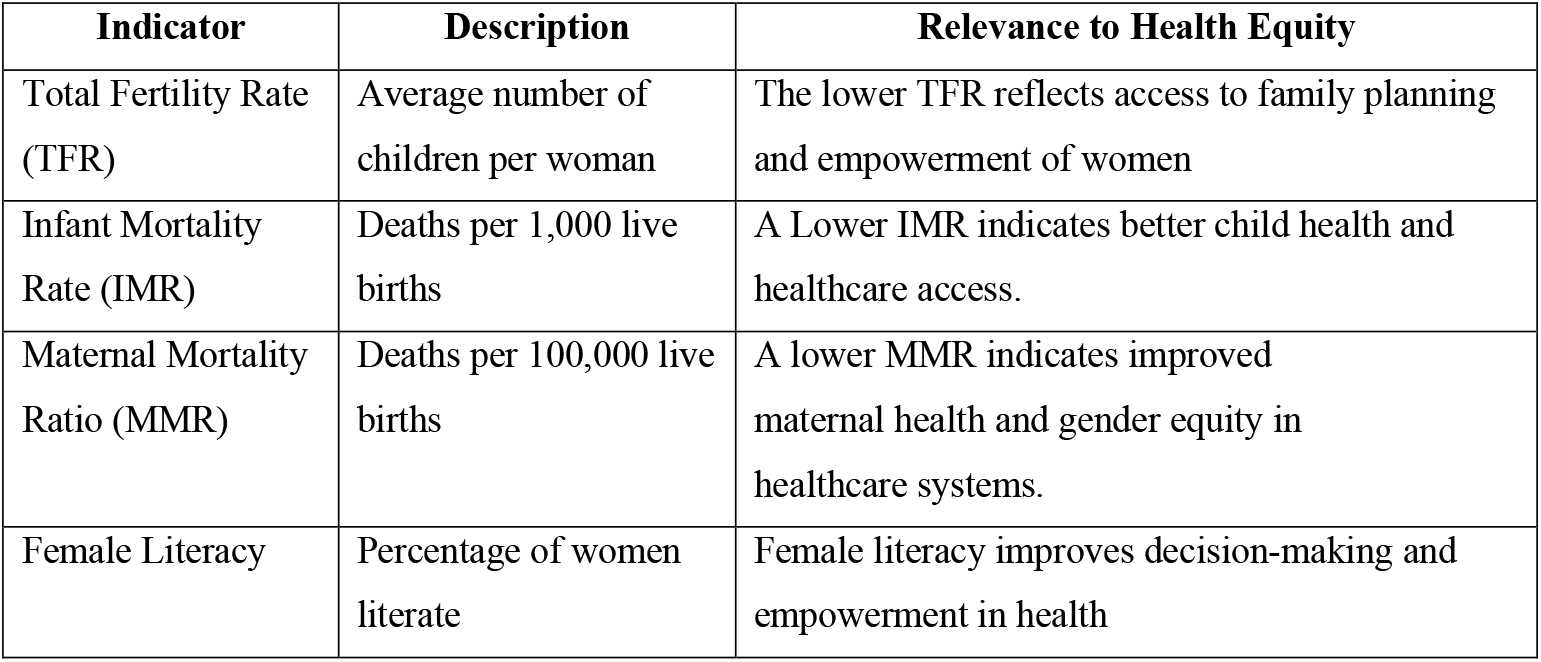
Health Indicators and Their Role in Delimitation.

| Indicator | Description | Relevance to Health Equity |
| --- | --- | --- |
| Total Fertility Rate (TFR) | Average number of children per woman | The lower TFR reflects access to family planning and empowerment of women |
| Infant Mortality Rate (IMR) | Deaths per 1,000 live births | A Lower IMR indicates better child health and healthcare access. |
| Maternal Mortality Ratio (MMR) | Deaths per 100,000 live births | A lower MMR indicates improved maternal health and gender equity in healthcare systems. |
| Female Literacy | Percentage of women literate | Female literacy improves decision-making and empowerment in health |

To create a unified health-weighted composite, all variables were normalised to a 0-1 scale using the formula:

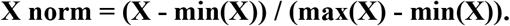

For IMR, MMR, and TFR, reverse scoring (X reverse = 1 - X norm) was applied so that higher normalised values represented better health performance, while population and female literacy were normalised directly. Principal Component Analysis (PCA) was conducted on the normalised metrics to derive a composite Health-Weighted Performance Score (HWPS). Diagnostics confirmed that the data was suitable for PCA: the correlation matrix showed strong, interpretable associations between metrics (for example, IMR and TFR, r = 0.83). The Kaiser-Meyer-Olkin (KMO) measure of sampling adequacy was 0.80. The Bartlett’s test of sphericity was significant (χ^2^ = 133.1, p < 0.001). The first principal component (PC1) explained 70.3% of the total variance. It was characterised by positive loadings for female literacy (0.477), TFR (0.506), IMR (0.488), and MMR (0.392), and a negative loading for population (-0.351). HWPS for each region was then calculated by rescaling PC1 to a 1-100 range using the formula:

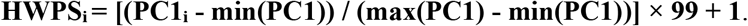

Pearson correlations supported the validity of HWPS: HWPS was strongly positively correlated with female literacy (r = 0.90, p < 0.0001) and negatively correlated with population size (r = -0.66, p < 0.0001), confirming that the score reflects health-related advantage rather than demographic weight. Seats were apportioned to each state and UT in proportion to their HWPS under three scenarios: current allocation (543 seats), a 10% expansion (597 seats), and a 15% expansion (625 seats). The initial raw entitlement for each state or UT was calculated as follows:

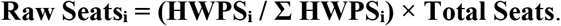

To ensure both fairness and political feasibility, two constraints were imposed in the final allocation: no region could have fewer seats than its 2019 seat count (the minimum safeguard), and (2) no region could have more seats than 110% (for 597 seats) or 115% (for 625 seats) of their 2019 seat count. The cap for each region was as follows:

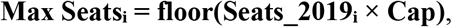

Where Cap = 1.10 or 1.15, as appropriate. Remaining seats after floors and caps were distributed by the largest fractional remainder method, up to the cap, until all seats were assigned.

To assess the robustness and stability of the model, 1,000 Monte Carlo simulations were conducted for each scenario, adding +/-10% Gaussian noise to all input metrics. Sensitivity analyses were performed by systematically increasing and decreasing each indicator by 20% to examine the impact on seat allocation. The consistency of results across all simulations and perturbations was used to evaluate the reliability of the health-weighted seat apportionment approach The analysis was conducted in R (version 4.3.2), utilising the packages *‘dplyr’* for data manipulation, ‘*scales’* for normalisation, ‘*psych’* for statistical diagnostics (KMO and Bartlett tests), *‘sf’* for spatial mapping, and *‘ggplot2’* for visualisation.

## Results

The Health-Weighted Performance Score (HWPS) ranged from 1 to 100, with high values concentrated in regions exhibiting low fertility and mortality and high female literacy, such as Kerala (88.9), Goa (89.4), Puducherry (97.0), and Lakshadweep (100.0). States with the largest populations and poorest health indicators, notably Bihar (1.0) and Uttar Pradesh (6.8), recorded the lowest HWPS. Across all three seat apportionment scenarios, like the current (543 seats), 10% expansion (597 seats), and 15% expansion (625 seats), no state or Union Territory lost seats due to the no-loss safeguard. Under a 10% expansion, 26 of 36 states and UTs gained at least one seat. States with higher HWPS, such as Kerala, Tamil Nadu, Maharashtra, and Delhi, saw clear gains: for example, Kerala increased from 20 (2019) to 22 (597 seats) and 23 (625 seats), while Uttar Pradesh and Bihar, despite their population size, received only modest increases.

Monte Carlo simulations (n = 1,000) demonstrated high stability: for each scenario, seat allocations for every region varied by no more than +/-1 seat in 95% of simulations. Sensitivity analyses confirmed robustness, with each indicator’s seat distributions stable under +/-20% perturbations. HWPS correlated strongly with female literacy (r = 0.90, p < 0.0001) and negatively with population (r = -0.66, p < 0.0001), validating its dual emphasis on health achievement and demographic context. Full allocations, HWPS, and simulation bounds for all scenarios are detailed in Table 3. The Health-Weighted Delimitation Scores of states and Union territories can be visualised through a choropleth map in Figure 1 and the scree plot in Figure 2.

**Figure 1:**
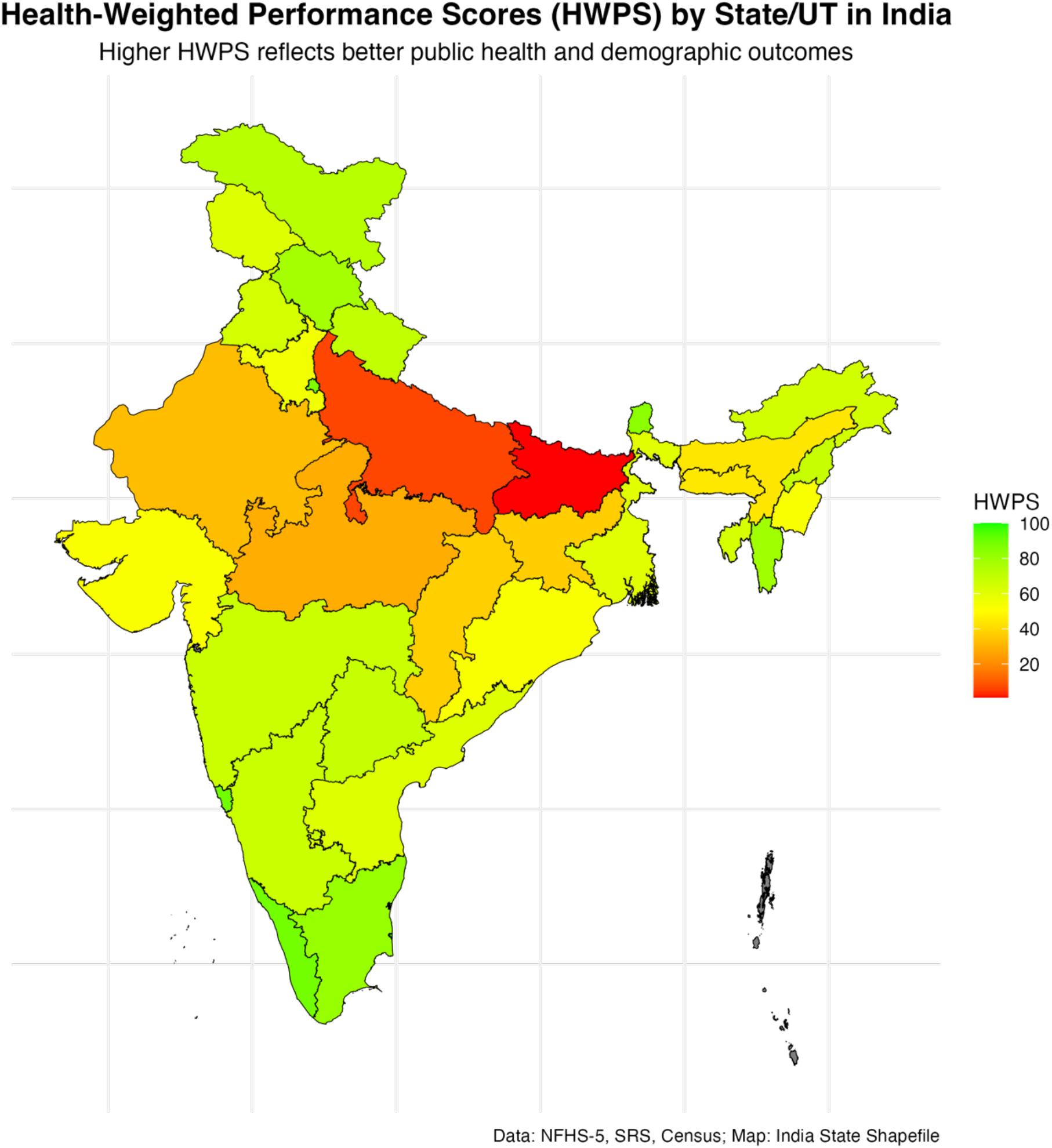

**Figure 2:**
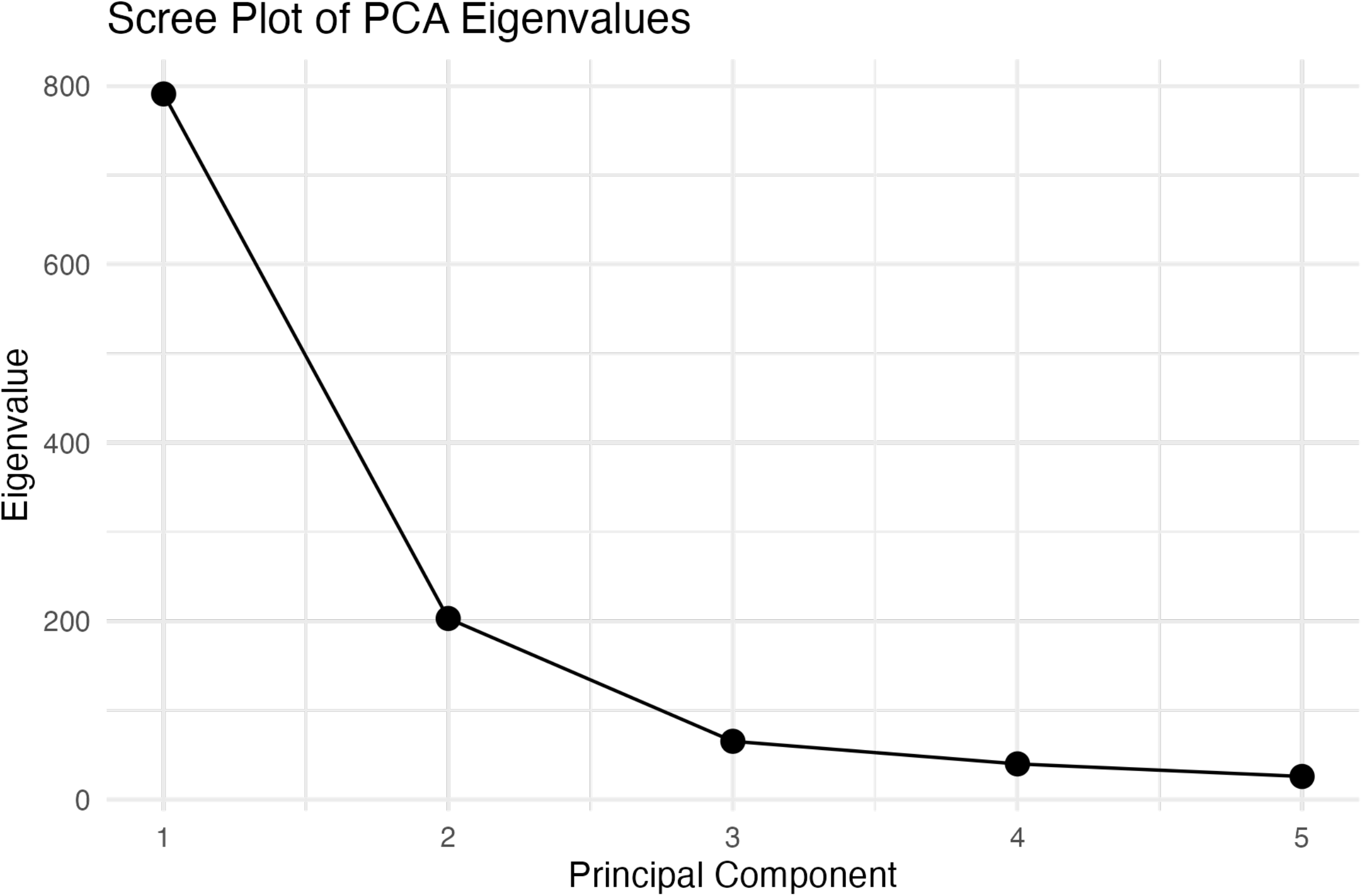

## Discussion

This nationwide simulation demonstrates the feasibility and policy relevance of health-weighted seat apportionment for India’s Lok Sabha. By integrating five key health indicators, total fertility rate (TFR), infant mortality rate (IMR), maternal mortality ratio (MMR), female literacy, and population into a Health-Weighted Performance Score (HWPS), the model aligns parliamentary representation with public health priorities. The first principal component captured 70.3% of the variance, with strong weights for maternal and infant health, female literacy, and fertility, while population plays a secondary role.

In the 10% expansion scenario (597 seats), the HWPS allocated additional seats to 26 out of 36 states and Union Territories, with a no-loss safeguard ensuring that no region falls below its 2019 baseline. High-performing states like Kerala, Tamil Nadu, and Maharashtra, with strong health and literacy metrics, have gained significantly, while populous states like Bihar and Uttar Pradesh have grown slightly. This trend has been maintained in the 15% expansion (625 seats), rewarding health progress without penalising structurally challenged regions and enhancing political feasibility. Robustness was confirmed through 1,000 Monte Carlo simulations (+/-10% Random Variation) and +/-20% sensitivity analyses, where seat allocation varied by a maximum of two seats and the no-loss safeguard prevented losses even at a population under-estimation of five per cent. This balances health incentives with institutional stability.

A National Task Force with Health Ministry and Election Commission members could pilot this idea. This reform advances the Delimitation Act 2002 and makes India the first to link seat allocation with health outcomes. This model offers an open-source, reproducible template for adding health equity to parliamentary representation for diverse federations. Figure 3 illustrates the framework for the implementation of Health-Weighted Delimitation, and Table 2 presents the Strengths, Weaknesses, Opportunities, Threats (SWOT) analysis.

**Figure 3:**
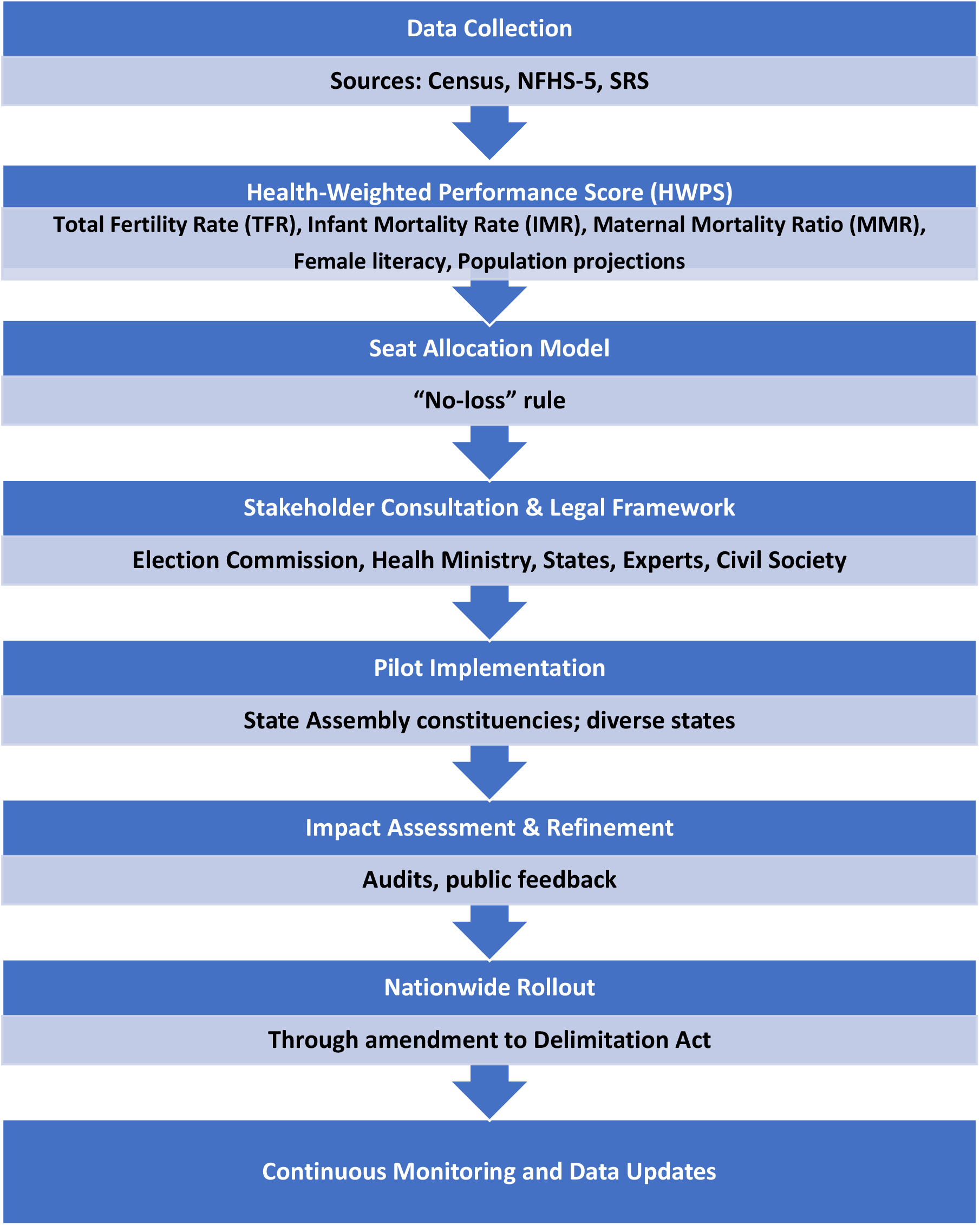
Proposed Health-Weighted Delimitation Framework.

**Table 2:** SWOT Analysis.

| Strengths | Weaknesses |
| --- | --- |
| Aligns representation with health equity | Political and constitutional hurdles |
| Encourages health investment | Requires robust data |
| Leverages existing data systems | Risk of metric politicisation |
| Opportunities | Threats |
| Global leadership in health-linked governance | Resistance from high-population states |
| Scalable pilot-to-national model | Fear of reduced voice for low-performing states |

### Challenges and Mitigation Strategies

The limitations of this model are the exclusion of vaccination rates and social determinants of health, such as sanitation, due to data gaps; these could be integrated in future models. Populous states like Uttar Pradesh and Bihar may resist lower HWPS-based influence. This can be mitigated by stakeholder involvement. Although politically necessary, the no-loss safeguard clause preserves the predominance of large-population states. Further reforms, such as fiscal transfers or more seats in the Upper House, may be recommended. Regional migration or fertility trends may be overlooked in the single population projections, which must be updated after the census. Independent audits and third-party verification using NFHS and SRS data are essential to avoid indicator gaming (e.g. inflated IMR or MMR). Transparent dashboards and awareness campaigns can help to increase public confidence.

## Conclusion

India’s demographic and health heterogeneity demands forward-thinking electoral reforms that explicitly integrate public health priorities. The health-weighted delimitation model described here, which synthesises total fertility rate, infant and maternal mortality, female literacy, and population size into a transparent composite score, offers a fair and evidence-based mechanism for parliamentary seat allocation. This model incentivises health system improvement by promoting equity and progress while protecting established representation. Implementing a pilot under the guidance of a National Task Force, alongside targeted legislative amendments to frameworks such as the Delimitation Act 2002, could enable its practical adoption. In doing so, India can lead by example, embedding health equity into democratic representation, a model other federations can follow.

## Data Availability

All data produced in the present work are contained in the manuscript

## Author and Contributor Statements

### Author Contributions

The author designed the study, conducted data collection and analysis, drafted the manuscript, and approved the final version for submission.

### Data Verification

The author accessed and verified all underlying data reported in the manuscript.

### Research in Context

#### Evidence before this study

Previous analyses of India’s demography and health disparities have shown significant regional differences in maternal and infant mortality, fertility, and female literacy based on sources such as the National Family Health Survey (NFHS) and Sample Registration System(SRS). However, no previous research has proposed or tested a composite health-related metric to guide the allocation of parliamentary seats, and current approaches rely almost exclusively on population size.

#### Added value of this study

This study introduces the Health-Weighted Performance Score (HWPS), a composite index derived from the five key health and demographic indicators by principal component analysis. This model, which apportions Lok Sabha seats in proportion to the HWPS while protecting existing minimum thresholds, demonstrates a feasible, equitable approach that directly integrates public health outcomes into political representation.

#### Implications of all the available evidence

The HWPS model provides a transparent and reliable way to link parliamentary representation with population and health outcomes. It encourages progress while ensuring support for disadvantaged regions. Using this approach could fix imbalances in representation and help leaders pay more attention to states that are falling behind. This idea could also be helpful for other federal democracies.

## Data Sharing Statement

All data used in this study are presented in Table 1. The R code for data analysis, including principal component analysis and seat allocation algorithms, is available upon request from the corresponding author (Dr. Karthik Balajee Laksham).

---

**Table 1:** Health-Weighted Delimitation Simulatio.

| State/UT | Population | IMR | MMR | TFR | Female<br>Literacy | HWPS | Seats<br>2019 | A | B | C | B<br>Lower | B<br>Upper | C<br>Lower | C<br>Upper |
| --- | --- | --- | --- | --- | --- | --- | --- | --- | --- | --- | --- | --- | --- | --- |
| Andhra Pradesh | 52883000 | 24 | 65 | 1.7 | 59.5 | 61.40 | 25 | 25 | 26 | 26 | 26 | 26 | 26 | 26 |
| Arunachal Pradesh | 1540000 | 13 | 100 | 1.9 | 59.6 | 64.85 | 2 | 2 | 2 | 2 | 2 | 2 | 2 | 2 |
| Assam | 35357000 | 32 | 195 | 1.9 | 67.3 | 43.71 | 14 | 14 | 15 | 15 | 15 | 15 | 15 | 15 |
| Bihar | 123082000 | 47 | 149 | 3 | 50.6 | 1.00 | 40 | 40 | 41 | 41 | 41 | 41 | 41 | 41 |
| Chhattisgarh | 29436000 | 38 | 137 | 2.2 | 60.2 | 37.43 | 11 | 11 | 12 | 12 | 12 | 12 | 12 | 12 |
| Goa | 1584000 | 10 | 45 | 1.5 | 73.4 | 89.37 | 2 | 2 | 2 | 2 | 2 | 2 | 2 | 2 |
| Gujarat | 68349000 | 31 | 87 | 2 | 66.8 | 52.02 | 26 | 26 | 27 | 27 | 27 | 27 | 27 | 27 |
| Haryana | 29688000 | 28 | 90 | 2.1 | 66.8 | 55.09 | 10 | 11 | 11 | 11 | 11 | 11 | 11 | 11 |
| Himachal Pradesh | 7614000 | 23 | 55 | 1.7 | 76.1 | 77.64 | 4 | 4 | 4 | 4 | 4 | 4 | 4 | 4 |
| Jharkhand | 37403000 | 34 | 103 | 2.4 | 56.2 | 36.93 | 14 | 14 | 15 | 15 | 15 | 15 | 15 | 15 |
| Karnataka | 67481000 | 22 | 68 | 1.8 | 72.9 | 67.41 | 28 | 28 | 29 | 29 | 29 | 29 | 29 | 29 |
| Kerala | 38740000 | 16 | 46 | 1.6 | 87.1 | 88.90 | 20 | 20 | 22 | 23 | 21 | 22 | 21 | 23 |
| Madhya Pradesh | 85358000 | 41 | 113 | 2.3 | 55.7 | 28.72 | 29 | 29 | 30 | 30 | 30 | 30 | 30 | 30 |
| Maharashtra | 123144000 | 22 | 60 | 1.7 | 75.5 | 66.84 | 48 | 48 | 49 | 49 | 49 | 49 | 49 | 49 |
| Manipur | 3211000 | 17 | 171 | 2.2 | 66.9 | 52.70 | 2 | 2 | 2 | 2 | 2 | 2 | 2 | 2 |
| Meghalaya | 3419000 | 22 | 188 | 2.2 | 61.5 | 44.25 | 2 | 2 | 2 | 2 | 2 | 2 | 2 | 2 |
| Mizoram | 1239000 | 12 | 98 | 1.7 | 74.6 | 78.85 | 1 | 1 | 1 | 1 | 1 | 1 | 1 | 1 |
| Nagaland | 2196000 | 14 | 105 | 1.9 | 66.9 | 68.14 | 1 | 1 | 1 | 1 | 1 | 1 | 1 | 1 |
| Odisha | 46364000 | 29 | 92 | 2 | 64.6 | 53.25 | 21 | 21 | 22 | 22 | 22 | 22 | 22 | 22 |
| Punjab | 32197000 | 27 | 71 | 1.9 | 71.7 | 64.56 | 13 | 14 | 14 | 14 | 14 | 14 | 14 | 14 |
| Rajasthan | 79716000 | 37 | 122 | 2.3 | 59.3 | 32.84 | 25 | 25 | 26 | 26 | 26 | 26 | 26 | 26 |
| Sikkim | 700000 | 15 | 70 | 1.7 | 79.9 | 83.71 | 1 | 1 | 1 | 1 | 1 | 1 | 1 | 1 |
| Tamil Nadu | 78530000 | 15 | 54 | 1.6 | 80.7 | 80.97 | 39 | 39 | 40 | 40 | 40 | 40 | 40 | 40 |
| Telangana | 39300000 | 24 | 63 | 1.7 | 68.6 | 68.54 | 17 | 17 | 18 | 19 | 18 | 18 | 18 | 19 |
| Tripura | 4167000 | 20 | 130 | 2 | 73.9 | 63.81 | 2 | 2 | 2 | 2 | 2 | 2 | 2 | 2 |
| Uttar Pradesh | 225071000 | 43 | 118 | 2.6 | 52.7 | 6.76 | 80 | 80 | 81 | 81 | 81 | 81 | 81 | 81 |
| Uttarakhand | 11229000 | 25 | 63 | 1.8 | 70.5 | 69.76 | 5 | 5 | 5 | 5 | 5 | 5 | 5 | 5 |
| West Bengal | 101116000 | 24 | 77 | 1.8 | 70 | 60.27 | 42 | 42 | 43 | 43 | 43 | 43 | 43 | 43 |
| Andaman and<br>Nicobar | 417000 | 11 | 50 | 1.5 | 76.7 | 90.33 | 1 | 1 | 1 | 1 | 1 | 1 | 1 | 1 |
| Chandigarh | 1239000 | 12 | 60 | 1.7 | 83.3 | 88.80 | 1 | 1 | 1 | 1 | 1 | 1 | 1 | 1 |
| Dadra and<br>Nagar Haveli | 678000 | 14 | 55 | 1.8 | 73.9 | 80.49 | 1 | 1 | 1 | 1 | 1 | 1 | 1 | 1 |
| Daman and<br>Diu | 343000 | 13 | 52 | 1.6 | 73.9 | 85.25 | 1 | 1 | 1 | 1 | 1 | 1 | 1 | 1 |
| Delhi | 19727000 | 17 | 43 | 1.6 | 81.4 | 86.81 | 7 | 7 | 7 | 8 | 7 | 7 | 8 | 8 |
| Jammu and<br>Kashmir | 6641000 | 20 | 90 | 2 | 62.3 | 61.12 | 6 | 6 | 6 | 6 | 6 | 6 | 6 | 6 |
| Ladakh | 351000 | 15 | 80 | 1.8 | 66.9 | 72.56 | 1 | 1 | 1 | 1 | 1 | 1 | 1 | 1 |
| Puducherry | 1629000 | 10 | 40 | 1.5 | 84.7 | 97.01 | 1 | 1 | 1 | 1 | 1 | 1 | 1 | 1 |
| Lakshadweep | 64473 | 12 | 40 | 1.4 | 88.2 | 100.00 | 1 | 1 | 1 | 1 | 1 | 1 | 1 | 1 |

UT- Union Territory, IMR-Infant Mortality Rate, MMR- Maternal Mortality Ratio, TFR- Total Fertility Rate, HWPS- Health Weighted Performance Score, *Population projected from Census 2011 using CAGR 1.2 %*, Seats 2019 - Current Lok Sabha seats, Scenario **A** : Seat apportionment using Health-Weighted Performance Score (HWPS) for a total of 543 Lok Sabha seats (current allocation), **B**: Apportionment with a 10% expansion, for a total of 563 seats, **C**: Apportionment with a 15% expansion, for a total of 566 seats.

